# Monthly excess mortality across counties in the United States during the Covid-19 pandemic, March 2020 to February 2022

**DOI:** 10.1101/2022.04.23.22274192

**Authors:** Eugenio Paglino, Dielle J. Lundberg, Zhenwei Zhou, Joe A. Wasserman, Rafeya Raquib, Anneliese N. Luck, Katherine Hempstead, Jacob Bor, Samuel H. Preston, Irma T. Elo, Andrew C. Stokes

**Author notes:** CORRESPONDING AUTHOR: Andrew C. Stokes, 801 Massachusetts Avenue, Crosstown Building 362, Boston, MA, 02118.

## Abstract

Excess mortality is the difference between expected and observed mortality in a given period and has emerged as a leading measure of the overall impact of the Covid-19 pandemic that is not biased by differences in testing or cause-of-death assignment. Spatially and temporally granular estimates of excess mortality are needed to understand which areas have been most impacted by the pandemic, evaluate exacerbating and mitigating factors, and inform response efforts, including allocating resources to affected communities. We estimated all-cause excess mortality for the United States from March 2020 through February 2022 by county and month using a Bayesian hierarchical model trained on data from 2015 to 2019. An estimated 1,159,580 excess deaths occurred during the first two years of the pandemic (first: 620,872; second: 538,708). Overall, excess mortality decreased in large metropolitan counties, but increased in nonmetro counties, between the first and second years of the pandemic. Despite the initial concentration of mortality in large metropolitan Northeast counties, beginning in February 2021, nonmetro South counties had the highest cumulative relative excess mortality. These results highlight the need for investments in rural health as the pandemic’s disproportionate impact on rural areas continues to grow.

## Introduction

The Covid-19 pandemic has had a substantial impact on mortality in the United States, leading to declines in life expectancy rarely observed since the end of World War II.^1,2^ Estimates of excess mortality, which compare observed deaths to those expected in the absence of the pandemic, suggest that the true death toll of the pandemic is much larger than indicated by the official Covid-19 deaths alone.^3–7^ Deaths attributable to the pandemic may have been assigned to causes other than Covid-19 for several reasons. Lack of access to testing in the community, combined with the inconsistent use of post-mortem testing for suspected cases, likely resulted in a large share of undiagnosed Covid-19 infections and deaths, especially early in the pandemic.^8–12^ Additionally, persons with comorbid conditions may have had their cause of death assigned to the comorbid condition rather than to Covid-19.^13^ Finally, excess deaths not assigned to Covid-19 may also reflect deaths indirectly related to the pandemic, including deaths associated with reductions in access to health care, hospital avoidance due to fear of Covid-19 infection, increases in drug overdoses, and economic hardship leading to housing and food insecurity.^14–20^

For these reasons, it is beneficial to use excess mortality as a measure of the pandemic’s impact, particularly when examining geographic patterns in mortality. Estimates of excess mortality are more comparable spatially than Covid-19-assigned deaths alone, because states use different procedures to assign Covid-19 deaths and local death investigation systems may have different policies and resources that affect assignment of Covid-19 deaths.^9,21^ Furthermore, because many Covid-19 deaths were not assigned to Covid-19 early in the pandemic, excess mortality is likely to provide a more accurate measure of the pandemic’s impact for purposes of resource allocation and evaluating health disparities.^7,22,23^ Thus, continued tracking of excess mortality across time and space helps to clarify the total impact of the pandemic, identify where its impacts have been greatest, and implement the most appropriate policy responses.

Prior studies of excess mortality in the United States have primarily focused on national and state-level estimates,^5,6^ but estimating the full impact of the Covid-19 pandemic at the county-level is necessary to understand finer-grained geographic patterns of excess mortality. Although the prior study generated predictions of excess mortality for 1,470 county sets for all months of 2020 combined,^4^ to the best of our knowledge, there are no estimates of excess mortality at the county-month level across the first two years of the pandemic. Additionally, expanding these estimates to the second year of the pandemic is critical because the geographic impact of the pandemic has changed markedly since the first year due to changing national and state-level policies, the availability of vaccines, and the emergence of new variants.

In the present study, we employ a Bayesian hierarchical model to estimate all-cause excess mortality by month for 3,127 counties for the period from March 2020 to February 2022. In addition to generating county-month level estimates of excess mortality, we examine spatial patterning of these estimates across Census divisions and large metros, medium/small metros, and nonmetro areas between the first and second year of the pandemic.

## Results

Across 3,127 counties in the U.S., 620,872 estimated excess deaths occurred during the first year of the pandemic (March 2020 to February 2021), and 538,708 estimated excess deaths occurred during the second year (March 2021 to February 2022). This equals a total of 1,159,580 excess deaths during the first two years of the pandemic.

### Geographic Patterns in Relative Excess Mortality

**Table 1** shows excess deaths and relative excess mortality across combinations of U.S. Census Divisions and metro-nonmetro areas during each pandemic year. In the entire United States, relative excess mortality decreased in large metros from 23% of expected deaths in the first year to 16% in the second year. Meanwhile, relative excess mortality in nonmetro areas increased from 20% in the first year to 23% in the second year. The decrease in relative excess mortality in large metro areas between the first and second year was particularly large in the Middle Atlantic (28% to 8%), New England (16% to 5%), and Pacific (26% to 16%) divisions. The increase in relative excess mortality in nonmetro areas was largest in the Pacific (7% to 21%), New England (5% to 13%), and Mountain (23% to 30%) divisions. The divisions that had the highest relative excess mortality in nonmetro areas during the second year were the South Atlantic (30%), Mountain (30%), West South Central (27%), and East South Central (26%) divisions.

**Table 1.** Excess Mortality by U.S. Census Division and Metro-Nonmetro Areas, March 2020 - February 2022

|  | Excess Deaths |  |  |  | Relative Excess |  |  |  |
| --- | --- | --- | --- | --- | --- | --- | --- | --- |
|  | Mar 2020 - Feb 2021 |  | Mar 2021 - Feb 2022 |  | Mar 2020 - Feb 2021 |  | Mar 2021 - Feb 2022 |  |
|  | Median | Posterior Interval (90%) | Median | Posterior Interval (90%) | Median | Posterior Interval (90%) | Median | Posterior Interval (90%) |
| **East North Central** |  |  |  |  |  |  |  |  |
| Large Metro | 47,615 | ( 25,819 - 66,511) | 37,648 | ( 7,050 - 64,008) | 0.209 | ( 0.103 - 0.318) | 0.165 | ( 0.027 - 0.318) |
| Medium or Small Metro | 23,092 | ( 10,831 - 34,143) | 23,248 | ( 5,615 - 38,283) | 0.177 | ( 0.076 - 0.285) | 0.178 | ( 0.038 - 0.330) |
| Non Metro | 17,182 | ( 7,474 - 25,568) | 19,488 | ( 6,037 - 31,203) | 0.171 | ( 0.068 - 0.277) | 0.193 | ( 0.053 - 0.350) |
| **Total** | 87,754 | ( 44,296 - 126,019) | 80,454 | ( 18,795 - 133,211) | 0.191 | ( 0.088 - 0.299) | 0.175 | ( 0.036 - 0.327) |
| **East South Central** |  |  |  |  |  |  |  |  |
| Large Metro | 11,855 | ( 6,462 - 16,539) | 12,395 | ( 4,718 - 18,980) | 0.212 | ( 0.105 - 0.322) | 0.220 | ( 0.074 - 0.381) |
| Medium or Small Metro | 15,704 | ( 8,465 - 22,191) | 17,256 | ( 6,610 - 26,259) | 0.204 | ( 0.100 - 0.314) | 0.222 | ( 0.075 - 0.382) |
| Non Metro | 18,450 | ( 10,921 - 24,947) | 20,188 | ( 9,416 - 29,267) | 0.237 | ( 0.128 - 0.349) | 0.258 | ( 0.106 - 0.423) |
| **Total** | 45,978 | ( 26,123 - 63,552) | 49,786 | ( 20,812 - 74,441) | 0.218 | ( 0.113 - 0.328) | 0.234 | ( 0.086 - 0.397) |
| **Middle Atlantic** |  |  |  |  |  |  |  |  |
| Large Metro | 73,092 | ( 48,366 - 94,719) | 21,354 | ( -13,410 - 51,515) | 0.283 | ( 0.171 - 0.400) | 0.083 | ( -0.046 - 0.226) |
| Medium or Small Metro | 14,526 | ( 5,905 - 21,796) | 10,870 | ( -1,407 - 21,203) | 0.163 | ( 0.061 - 0.267) | 0.122 | ( -0.014 - 0.269) |
| Non Metro | 5,404 | ( 2,322 - 8,091) | 7,044 | ( 2,704 - 10,745) | 0.166 | ( 0.065 - 0.270) | 0.216 | ( 0.073 - 0.372) |
| **Total** | 92,990 | ( 56,440 - 124,749) | 39,390 | ( -11,962 - 83,717) | 0.245 | ( 0.135 - 0.358) | 0.104 | ( -0.028 - 0.249) |
| **Mountain** |  |  |  |  |  |  |  |  |
| Large Metro | 22,560 | ( 14,456 - 29,674) | 21,603 | ( 9,756 - 31,528) | 0.267 | ( 0.156 - 0.384) | 0.254 | ( 0.101 - 0.419) |
| Medium or Small Metro | 15,580 | ( 8,494 - 21,839) | 17,856 | ( 7,726 - 26,423) | 0.210 | ( 0.104 - 0.321) | 0.237 | ( 0.091 - 0.397) |
| Non Metro | 8,460 | ( 4,968 - 11,627) | 11,242 | ( 5,892 - 15,570) | 0.225 | ( 0.121 - 0.337) | 0.296 | ( 0.136 - 0.462) |
| **Total** | 46,534 | ( 27,880 - 63,092) | 50,537 | ( 23,686 - 73,493) | 0.237 | ( 0.130 - 0.351) | 0.254 | ( 0.105 - 0.418) |
| **New England** |  |  |  |  |  |  |  |  |
| Large Metro | 10,970 | ( 4,603 - 16,564) | 3,110 | ( -6,025 - 10,822) | 0.164 | ( 0.063 - 0.270) | 0.046 | ( -0.079 - 0.182) |
| Medium or Small Metro | 7,280 | ( 2,329 - 11,565) | 3,392 | ( -3,746 - 9,343) | 0.141 | ( 0.041 - 0.245) | 0.065 | ( -0.063 - 0.203) |
| Non Metro | 1,036 | ( -827 - 2,702) | 2,488 | ( -263 - 4,748) | 0.053 | ( -0.038 - 0.150) | 0.125 | ( -0.012 - 0.270) |
| **Total** | 19,206 | ( 6,157 - 30,859) | 8,981 | ( -10,084 - 24,924) | 0.139 | ( 0.041 - 0.244) | 0.065 | ( -0.064 - 0.203) |
| **Pacific** |  |  |  |  |  |  |  |  |
| Large Metro | 63,256 | ( 39,598 - 83,818) | 38,569 | ( 4,950 - 66,796) | 0.257 | ( 0.147 - 0.372) | 0.157 | ( 0.018 - 0.308) |
| Medium or Small Metro | 17,046 | ( 6,465 - 26,432) | 20,888 | ( 5,751 - 33,902) | 0.152 | ( 0.053 - 0.257) | 0.185 | ( 0.045 - 0.340) |
| Non Metro | 2,070 | ( -815 - 4,542) | 6,324 | ( 2,162 - 9,741) | 0.069 | ( -0.025 - 0.165) | 0.209 | ( 0.063 - 0.362) |
| **Total** | 82,420 | ( 45,325 - 114,878) | 65,735 | ( 13,048 - 110,745) | 0.212 | ( 0.107 - 0.323) | 0.169 | ( 0.030 - 0.323) |
| **South Atlantic** |  |  |  |  |  |  |  |  |
| Large Metro | 54,882 | ( 27,730 - 79,127) | 56,126 | ( 17,067 - 89,532) | 0.191 | ( 0.088 - 0.301) | 0.194 | ( 0.052 - 0.351) |
| Medium or Small Metro | 44,112 | ( 22,014 - 63,596) | 50,446 | ( 18,369 - 77,446) | 0.190 | ( 0.086 - 0.298) | 0.214 | ( 0.069 - 0.372) |
| Non Metro | 22,575 | ( 14,187 - 29,897) | 26,516 | ( 14,290 - 36,817) | 0.254 | ( 0.146 - 0.366) | 0.298 | ( 0.141 - 0.468) |
| **Total** | 121,399 | ( 64,366 - 172,700) | 132,882 | ( 50,027 - 203,515) | 0.200 | ( 0.097 - 0.310) | 0.217 | ( 0.072 - 0.375) |
| **West North Central** |  |  |  |  |  |  |  |  |
| Large Metro | 11,422 | ( 5,277 - 16,749) | 9,900 | ( 1,024 - 17,369) | 0.178 | ( 0.075 - 0.284) | 0.153 | ( 0.014 - 0.304) |
| Medium or Small Metro | 10,810 | ( 4,780 - 15,986) | 9,114 | ( 620 - 16,292) | 0.174 | ( 0.070 - 0.280) | 0.146 | ( 0.009 - 0.294) |
| Non Metro | 12,152 | ( 5,188 - 18,648) | 11,184 | ( 1,279 - 19,777) | 0.163 | ( 0.064 - 0.275) | 0.150 | ( 0.015 - 0.301) |
| **Total** | 34,253 | ( 15,484 - 51,120) | 30,142 | ( 2,973 - 53,340) | 0.170 | ( 0.070 - 0.278) | 0.150 | ( 0.013 - 0.299) |
| **West South Central** |  |  |  |  |  |  |  |  |
| Large Metro | 36,592 | ( 22,404 - 48,792) | 33,477 | ( 13,086 - 50,997) | 0.247 | ( 0.138 - 0.359) | 0.223 | ( 0.077 - 0.385) |
| Medium or Small Metro | 32,270 | ( 21,799 - 41,142) | 28,219 | ( 13,749 - 40,632) | 0.302 | ( 0.186 - 0.420) | 0.263 | ( 0.113 - 0.428) |
| Non Metro | 20,482 | ( 13,550 - 26,694) | 19,558 | ( 9,367 - 28,017) | 0.279 | ( 0.169 - 0.397) | 0.266 | ( 0.112 - 0.431) |
| **Total** | 89,258 | ( 57,879 - 116,753) | 81,377 | ( 36,448 - 119,338) | 0.272 | ( 0.161 - 0.388) | 0.246 | ( 0.097 - 0.408) |
| **United States** |  |  |  |  |  |  |  |  |
| Large Metro | 332,306 | (196,288 - 451,610) | 233,418 | ( 39,476 - 400,509) | 0.231 | ( 0.125 - 0.342) | 0.162 | ( 0.024 - 0.314) |
| Medium or Small Metro | 180,034 | ( 91,465 - 259,254) | 181,067 | ( 54,096 - 290,287) | 0.192 | ( 0.089 - 0.302) | 0.192 | ( 0.051 - 0.348) |
| Non Metro | 107,666 | ( 57,214 - 152,520) | 123,794 | ( 51,623 - 186,085) | 0.201 | ( 0.098 - 0.311) | 0.231 | ( 0.085 - 0.392) |
| **Total** | 620,872 | (345,616 - 863,761) | 538,708 | (145,084 - 876,551) | 0.213 | ( 0.108 - 0.324) | 0.184 | ( 0.044 - 0.339) |

**Notes:** Relative excess is the ratio of excess deaths to expected deaths, indicating the proportion increase in observed deaths compared to expected deaths during a period.

**Figure 1** shows the evolution of relative excess mortality across four mortality peaks during the pandemic across U.S. counties. These maps show the extent of excess mortality and demonstrate how excess mortality shifted from coastal regions early in the pandemic into the rest of the country as the pandemic progressed. During the Delta peak, excess mortality became more concentrated in the South and Mountain West, while the spatial distribution of excess mortality during the Omicron peak was less geographically consistent. **Figure 2** shows the probability of counties having any positive relative excess mortality across four mortality peaks during the pandemic, demonstrating that during each wave of the pandemic observed mortality fell above the range of values we would have expected in the absence of the COVID-19 pandemic.

**Figure 1.**
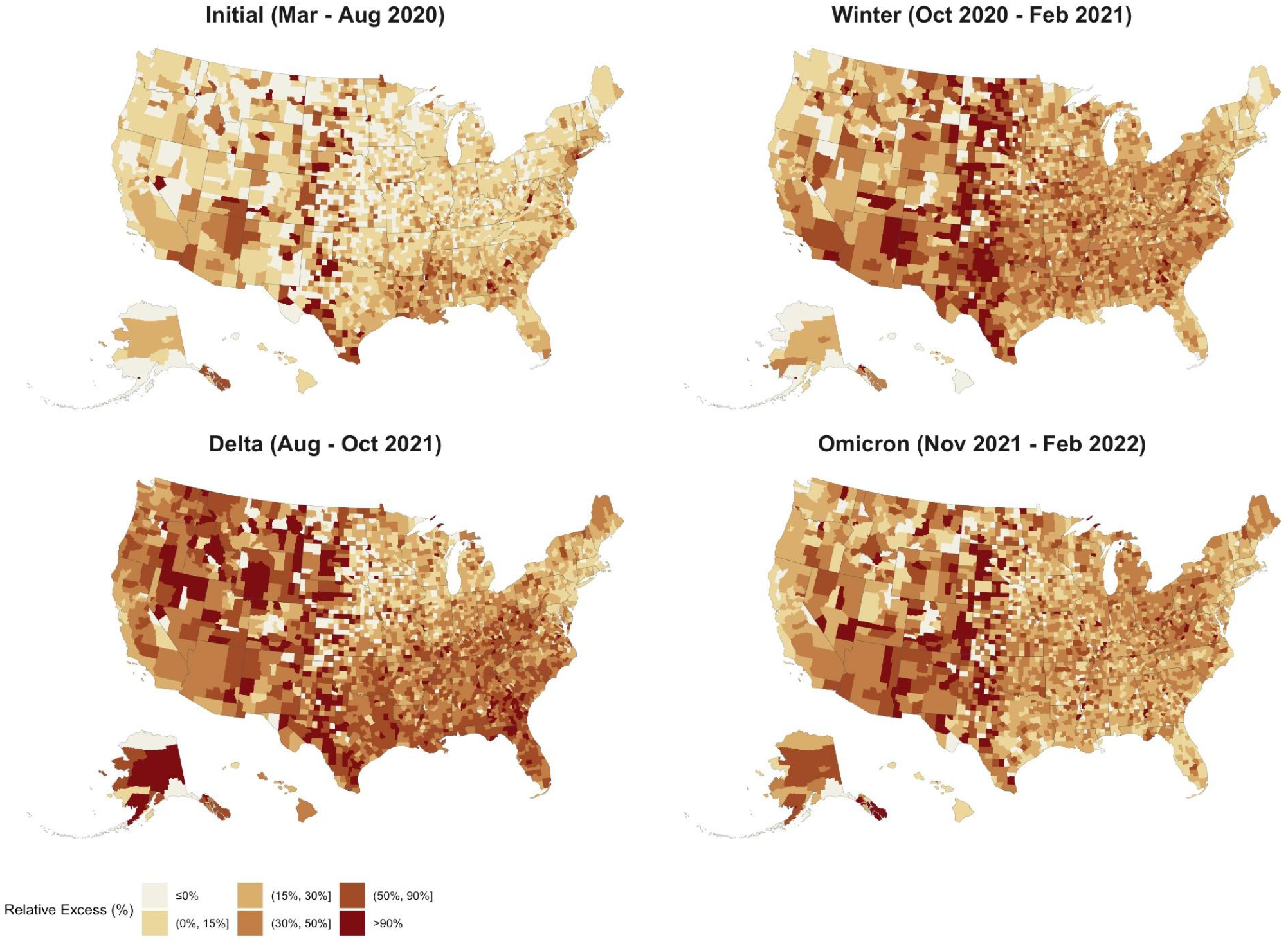
Relative Excess Mortality across U.S. Counties during 4 Mortality Peaks, March 2020 - February 2022 Notes: Each county in the map is colored according to its relative excess mortality (the ratio of excess deaths over expected deaths). Each of the four maps refers to one of the four peak periods of the pandemic, months of particularly high excess mortality. Category cutoffs are the 10th, 30th, 60th, 80th and 95th percentiles rounded to the nearest 5% relative excess.

**Figure 2.**
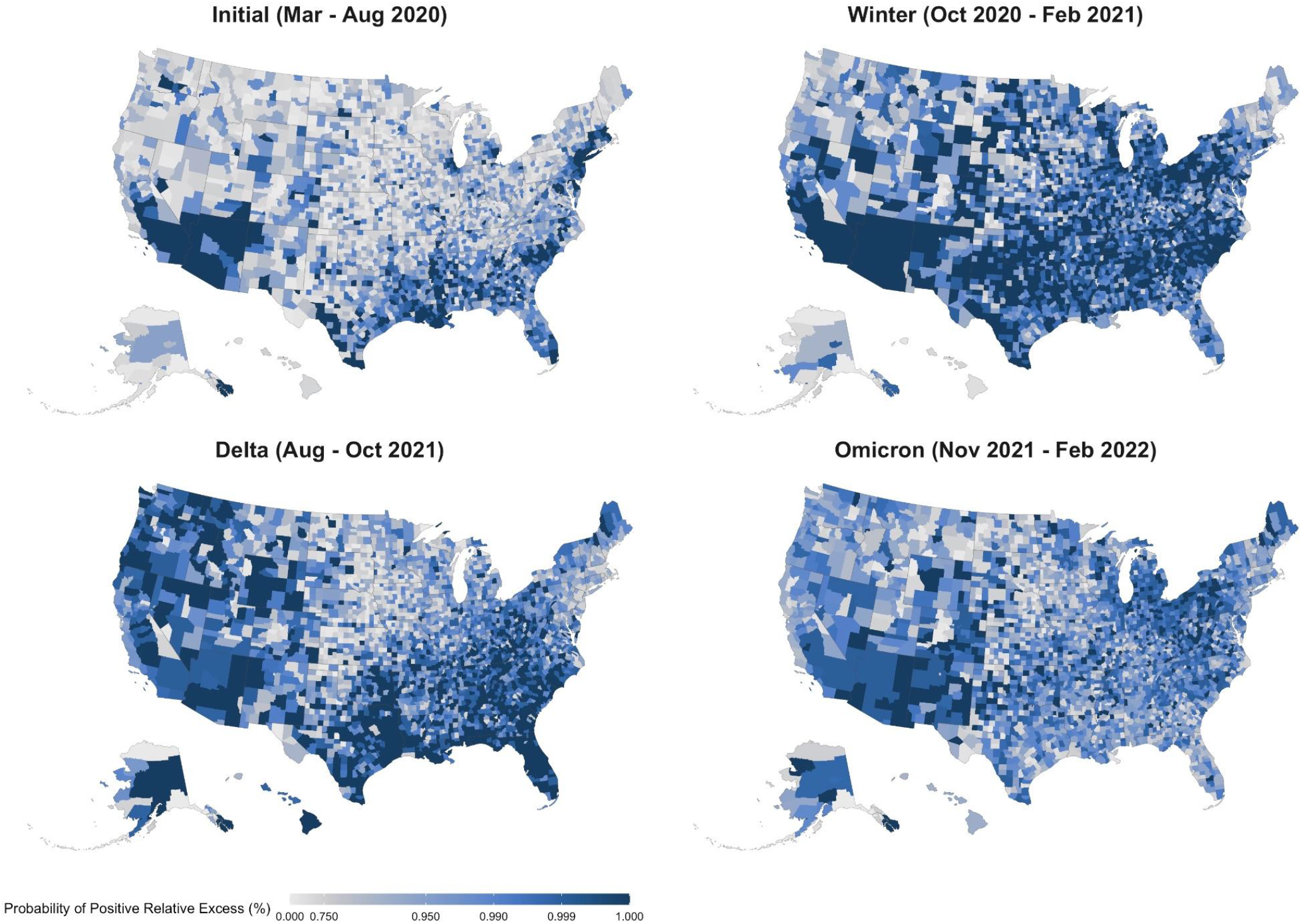
Probability of Positive Excess Mortality across U.S. Counties during 4 Mortality Peaks, March 2020 - February 2022 Notes: Each county in the map is colored according to the posterior probability that the observed death count is higher than the expected one. We highlight counties where the probability of positive excess mortality is higher than 0.75. The four maps refer to the four peak periods of the pandemic, months of particularly high excess mortality.

### Temporal Trends in Relative Excess Mortality

Throughout the pandemic, national trends in excess mortality reflect the aggregation of heterogeneous trends across disparate census regions and metro/nonmetro areas. To explore this pattern, **Figure 3** shows temporal trends in relative excess mortality across combinations of U.S. Census division and metro-nonmetro categories. The initial peak in excess mortality nationally was mostly driven by high excess mortality in large metro areas within the Middle Atlantic region. In contrast, the Winter peak spared this region and affected counties across the metro-nonmetro continuum in other regions of the country. As the pandemic progressed, there was a higher degree of concordance in temporal patterns across regions, which was especially evident during Delta and Omicron. **Figure 4** further illustrates differences in the geography of the pandemic between the first and second year by directly comparing relative excess mortality in the two years across divisions and metro-nonmetro areas. Large metro counties predominantly had greater relative excess mortality in the first year of the pandemic than they did in the second year. In contrast, nonmetro counties were more likely to have greater relative excess mortality in the second year as compared with the first year. This pattern is indicative of the emergence of a rural disadvantage in the second year of the pandemic.

**Figure 3.**
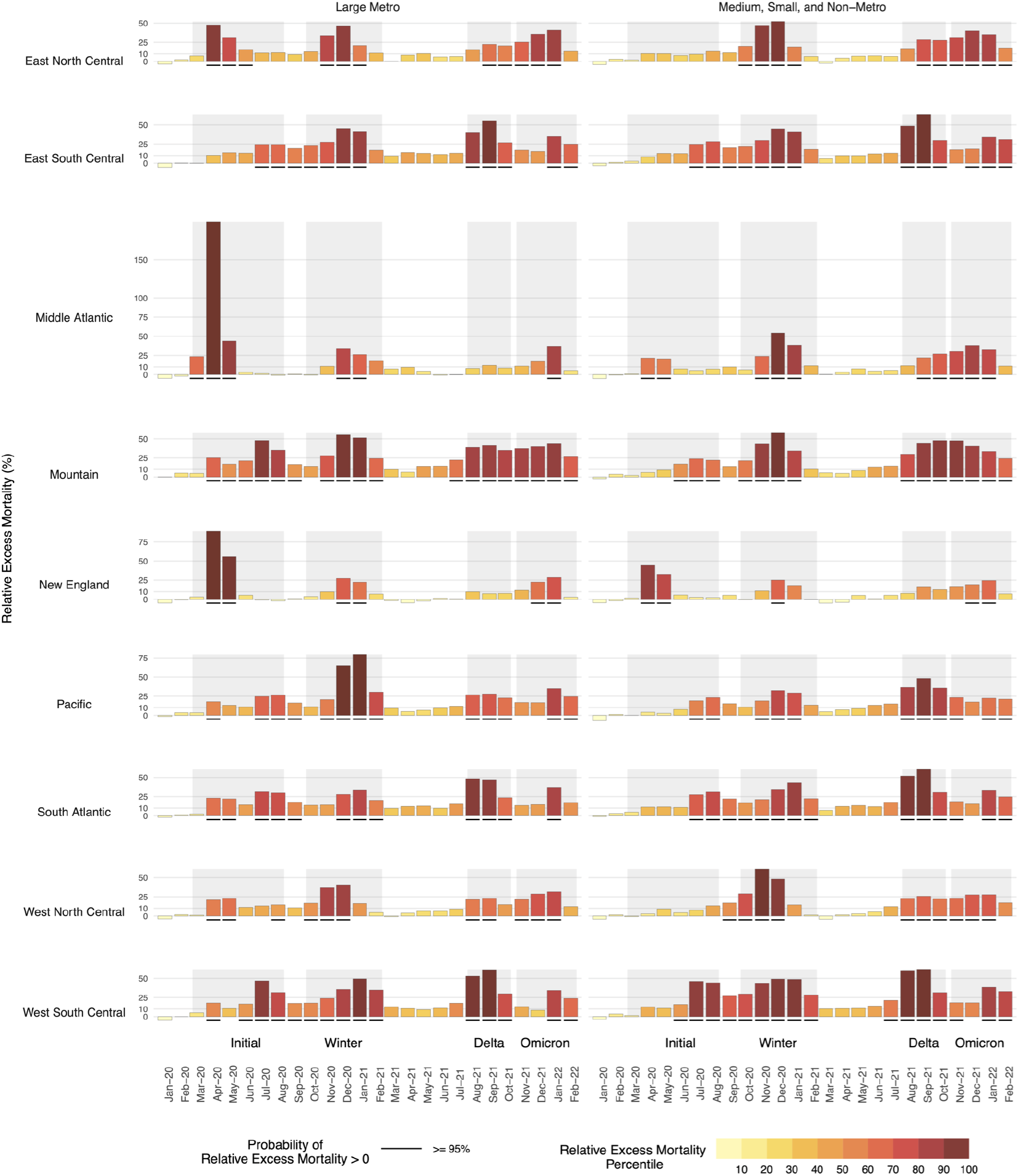
Temporal Trends in Relative Excess Mortality by Census Division and Metro-Nonmetro Status, March 2020 - February 2022 Notes: The large metro category includes large central metros and large fringe metros. All non large metro counties are classified as medium, small, and non-metro. The shaded intervals behind the bars separate the different waves of the COVID-19 pandemic as follow: Initial Wave (Mar 2020 - Aug 2020), Winter Peak (Oct 2020 - Feb 2021), Delta (Aug 2021 - Oct 2021), Omicron (Nov 2021 - Feb 2022). The height of each bar reflects excess deaths as a proportion of expected deaths. The color of the bars reflects each Division-month position (percentile) in the overall distribution of relative excess. Black, solid segments below the bars indicate units for which the posterior probability of positive excess mortality is above 95%.

**Figure 4.**
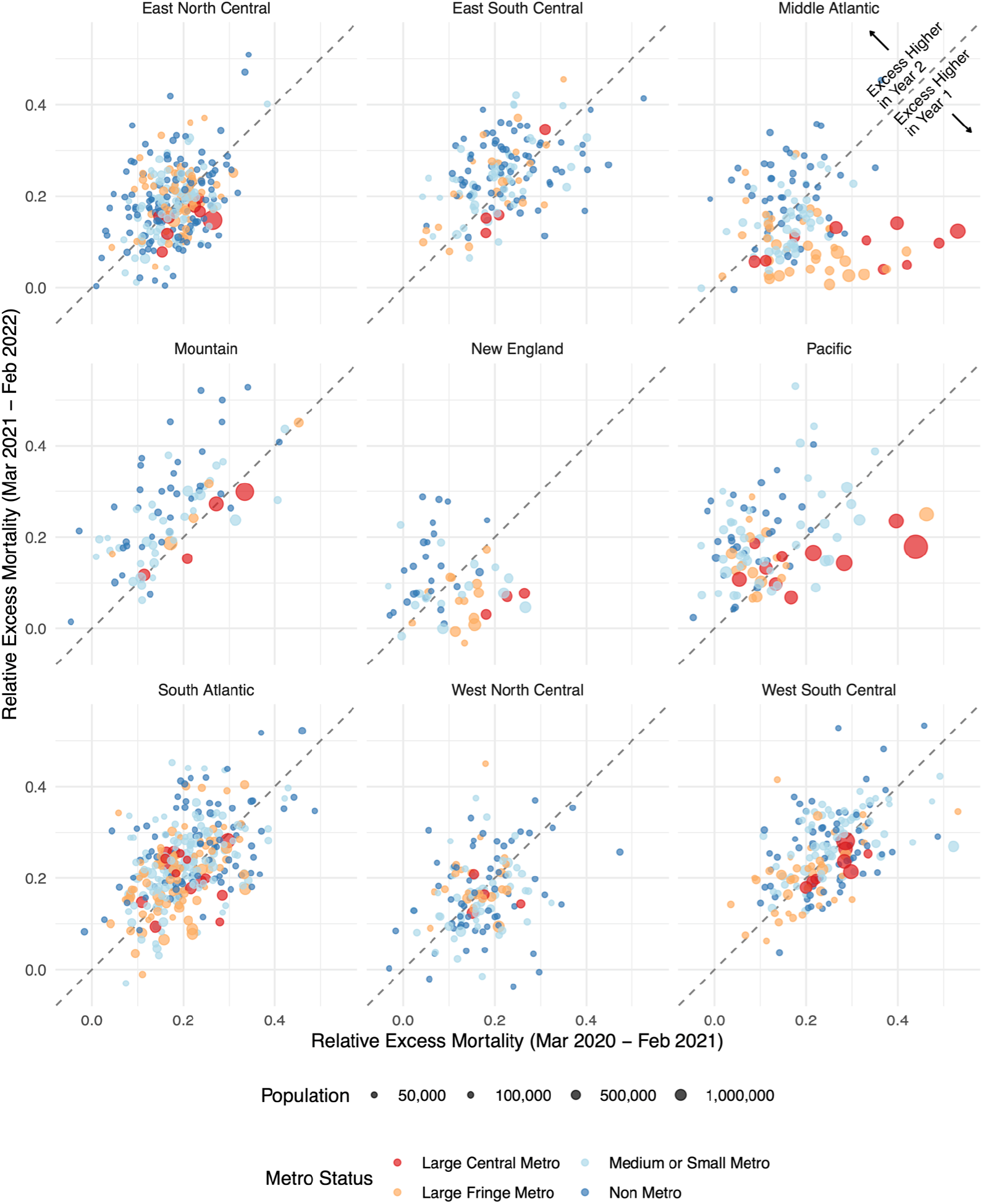
A Comparison of Relative Excess Mortality by Pandemic Year, March 2020 - February 2022 Notes: Each point in the graph represents a county and reflects its relative excess mortality from March 2020 to February 2021 (horizontal axis) and its relative excess mortality from March 2021 to February 2022 (vertical axis). We excluded counties with less than 30,000 residents to make the relationship between the two variables clearer. The 45 degrees line separates the plot into two parts. Points above the line saw higher mortality in the second year of the pandemic compared to the first one. Points falling below the line saw instead a decrease in mortality in the second year compared to the first.

### Cumulative Trends in Relative Excess Mortality

**Figure 5** examines relative excess mortality in cumulative terms for combinations of Census Region and metropolitan status. In the initial months of the pandemic, large metro areas in the Northeast experienced exceptionally high relative excess mortality. However, by February 2021, cumulative relative excess mortality in the non-metro South had exceeded cumulative relative excess mortality in large metro areas in the Northeast. Cumulative relative excess mortality remained higher in the non-metro South than in any other region through the end of February 2022. By the end of February 2022, cumulative relative excess mortality was lower in metro and nonmetro areas in the Northeast than in metro and nonmetro areas in the South, West, and Midwest. The regions with the highest cumulative relative excess mortality at the end of February 2022 were nonmetro areas in the South, medium and small metro areas in the South, large metros in the West, nonmetro areas in the West, and large metros in the South.

**Figure 5.**
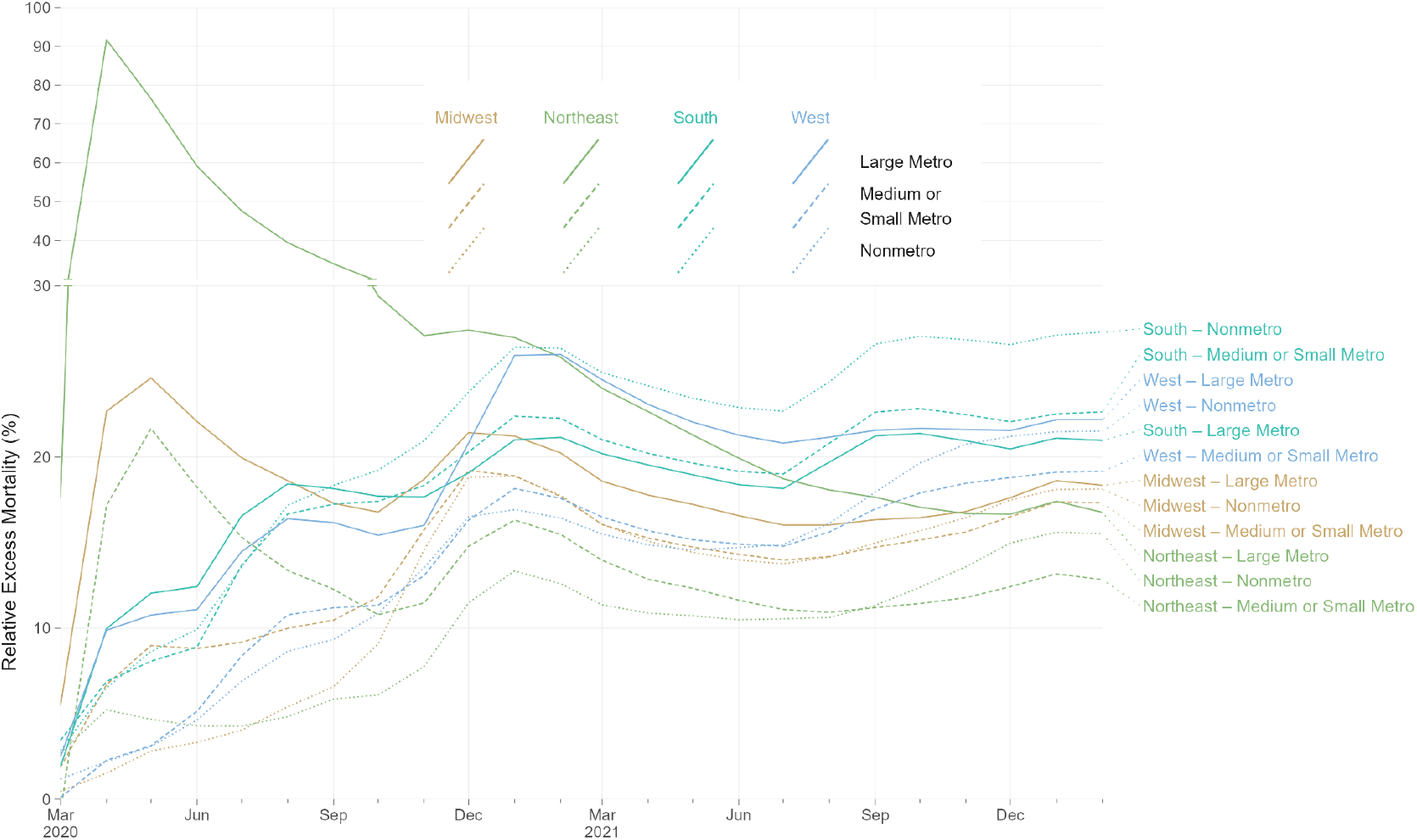
Rolling Cumulative Relative Excess Mortality by Census Region and Metro Category, March 2020 - February 2022 Notes: Each line represents the rolling cumulative excess mortality for one combination metropolitan/nonmetropolitan area categories and one census region. “Large Metro” includes large central and large fringe metropolitan areas. Each census region is represented by a different line color: light brown for Midwest, green for Northeast, aquamarine for South, and light blue for West. Each metropolitan category is represented by a different line type: solid for Large Metro, dashed for Medium or Small Metro, and dotted for Nonmetro. The y-axis for relative excess mortality above 30% is compressed vertically. Rolling cumulative relative excess mortality is calculated as the sum of excess deaths divided by the sum of expected deaths for all months from March, 2020, through a given month. For example, values for February, 2022, reflect total excess deaths for 24 months of the pandemic, from March, 2020, through February, 2022. Decreasing cumulative relative excess mortality indicates months with relative excess mortality below-average to date, for a given combination of census region and metro category. Increasing cumulative relative excess mortality indicates months with relative excess mortality above-average to date, for a given combination of census region and metro category.

### County-Level Trends in Relative Excess Mortality

An emerging rural disadvantage is also visible when examining temporal trends for individual counties. **Figure 6** shows temporal trends in relative excess mortality for the most populous counties among large metro and nonmetro counties. Among large metro counties, relative excess mortality was especially high in Northeastern counties in the early pandemic and in California counties during the Winter peak. In nonmetro counties, marked increases in mortality were observed during the second year of the pandemic, especially during the Delta peak. **Figure 7** explores changes in excess mortality between the first and second year of the pandemic among the most populous counties in each metro-nonmetro category. In the most populous counties in large metro areas, substantial declines in excess mortality were observed between the first and second year. For counties in nonmetro areas, the opposite pattern was observed. These areas were generally spared in the first year, after which they experienced high mortality in the second year. **Figure 8** displays temporal trends for each county alongside state trend lines. This figure reveals substantial variation in temporal trends in relative excess mortality across states along with substantial variation in relative excess mortality trends within states.

**Figure 6.**
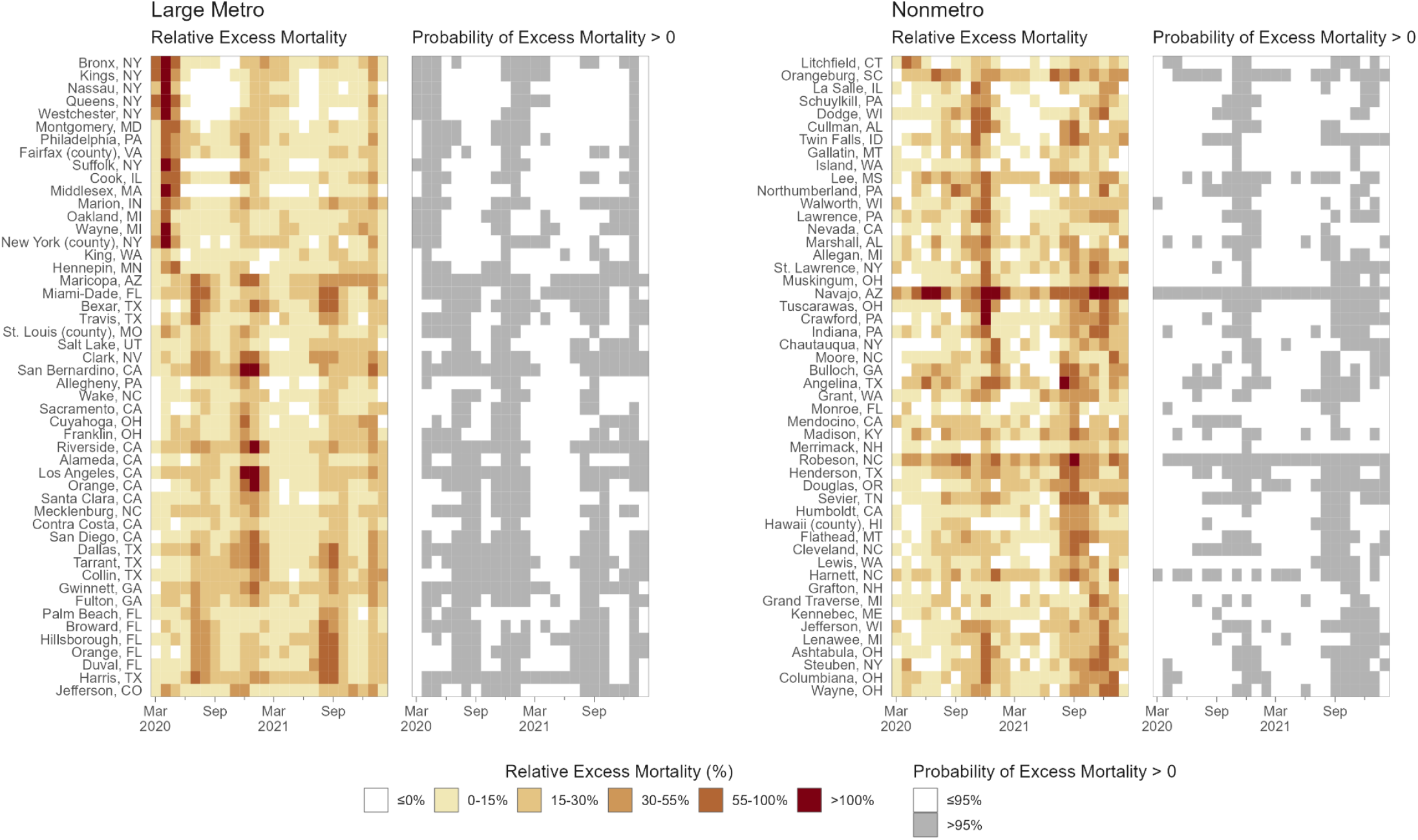
Spatial-Temporal Trends in Relative Excess Mortality Among the Most Populous Counties by Metro-Nonmetro Category, March 2020 - February 2022 Notes: Each cell in the four heatmaps represents a county-month. In the shaded heatmaps colored from white to dark red, darker and redder colors indicate higher relative excess mortality. In the white-and-gray heatmaps, gray cells indicate county-months with a greater than 90% probability of positive excess mortality. Counties in each pair of heatmaps are the 50 most populous within two categories of metropolitan and nonmetropolitan areas. “Large Metro” includes large central and large fringe metropolitan areas. Within each pair of heatmaps, counties were sorted vertically based on the occurrence of the highest peak of excess deaths. Counties at the tops of the heatmaps thus had their month of highest relative excess earlier in the pandemic. In contrast, those at the bottom had their highest peak or relative excess later in the pandemic.

**Figure 7.**
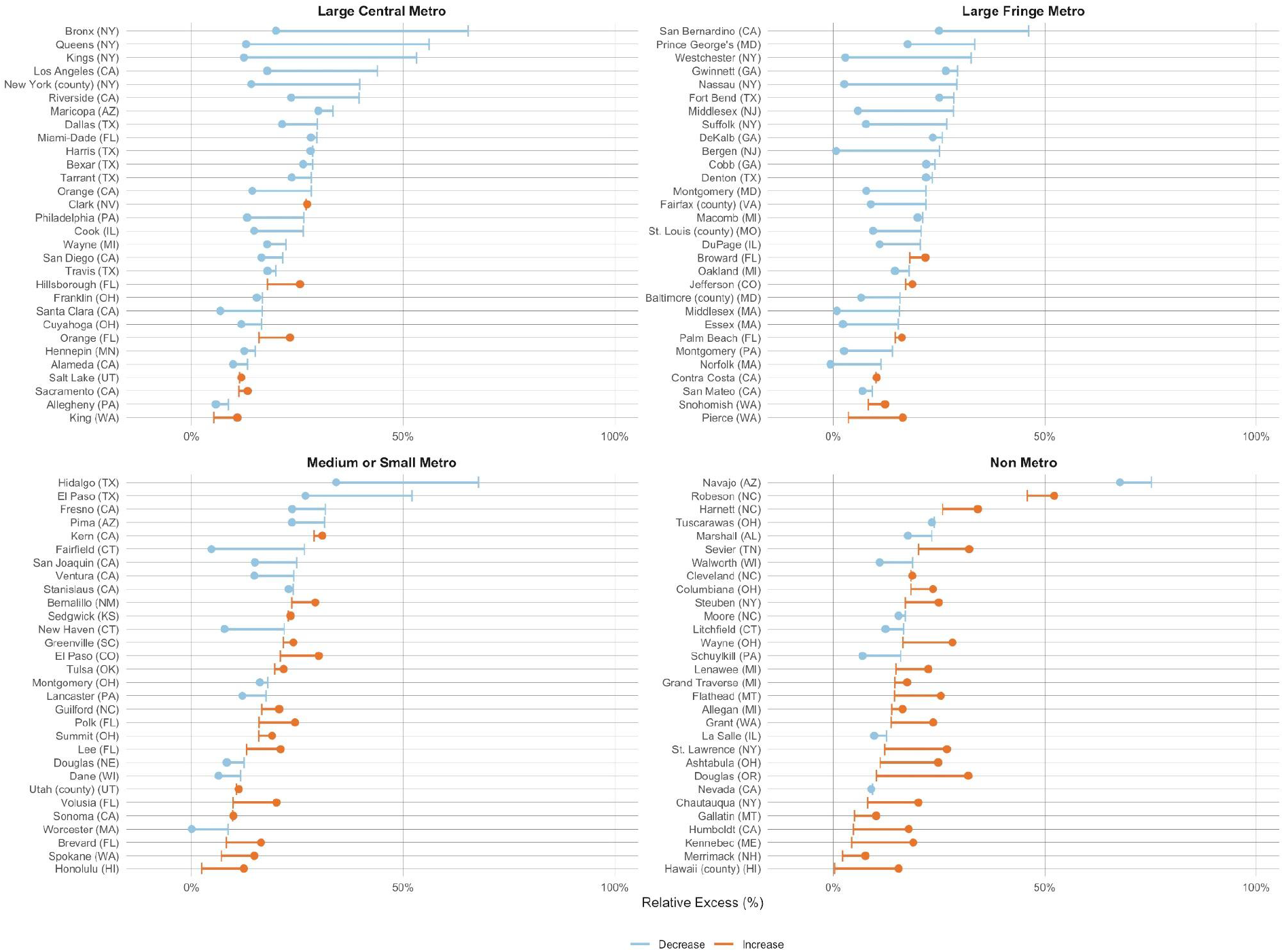
Change in Relative Excess Mortality between the First and Second Pandemic Years In the Most Populous Counties by Metro-Nonmetro Category, March 2020 - February 2022 Notes: Each line in the four graphs represents a county. For each line, the vertical segment reflects relative excess in the first year of the pandemic (Mar 2020 - Feb 2021), while the dot indicates relative excess in the second year of the pandemic (Mar 2021 - Feb 2022). The color of the line distinguishes between counties that saw a decline in relative excess (blue lines), and those that saw an increase (orange lines). The 30 most populous counties were selected for each metro category.

**Figure 8.**
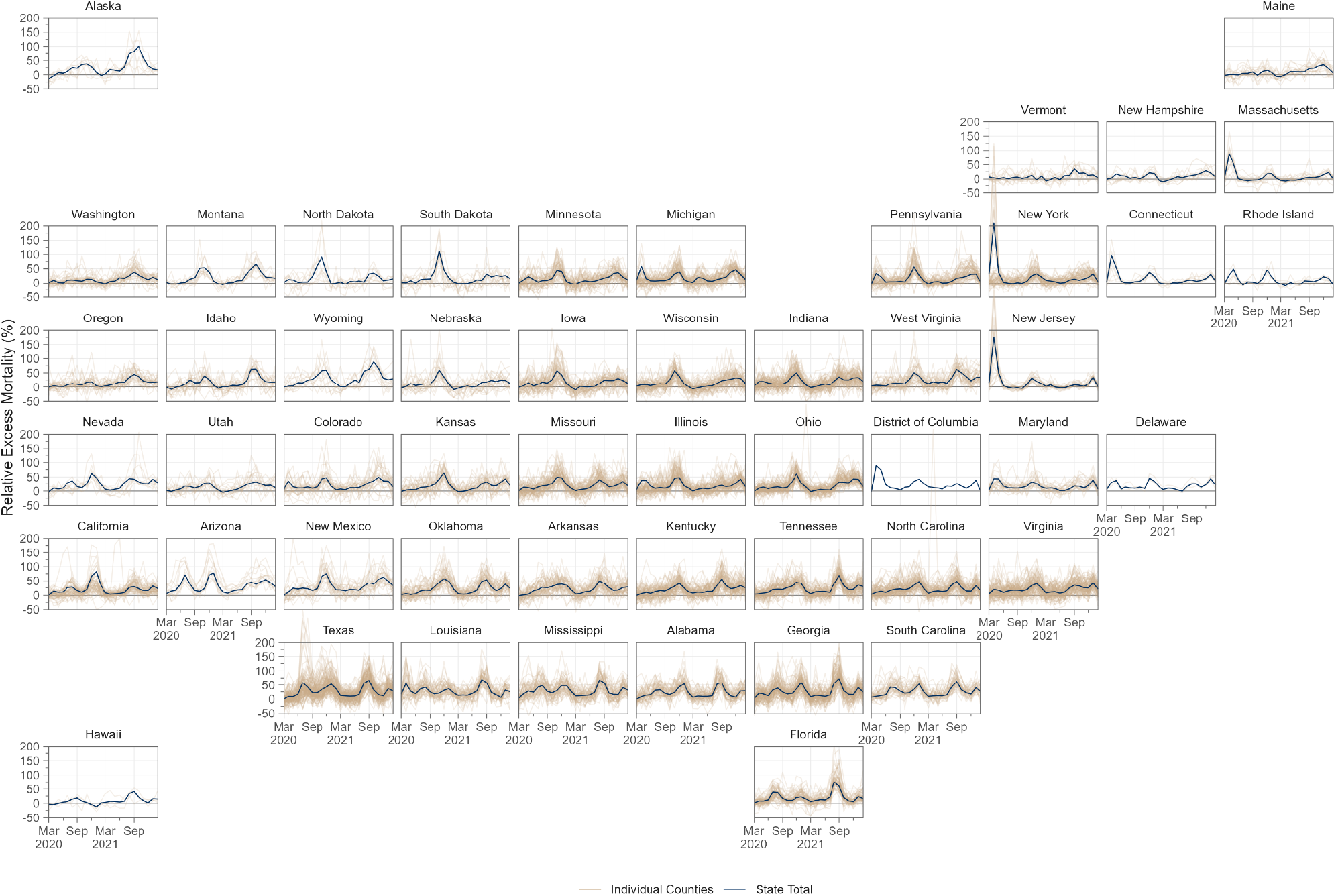
Temporal Trends in Relative Excess Mortality across Counties in Each State, March 2020 to February 2022 Notes: Each plot presents data for a different state, position in the figure reflects the approximate geographical position of each state within the US. State level monthly time series of relative excess are denoted by solid dark blue lines. Each light brown transparent line represents the monthly time series for one county in that state. Counties with greater than 15,000 population in all states in our sample are depicted.

## Discussion

This study produced monthly estimates of excess mortality for 3,127 counties in the U.S. from March 2020 through February 2022, identifying 1,159,580 excess deaths during the first two years of the pandemic. Between the first and second year of the pandemic, relative excess mortality decreased in large metro areas and increased in nonmetro areas. The increases in mortality in nonmetro areas occurred most markedly during the Delta wave of the pandemic. By the end of February 2022, nonmetro and medium and small metro areas in the South had the highest cumulative relative excess mortality, surpassing large metro areas in the Northeast and other areas that were affected heavily in the early pandemic.

Prior studies of excess mortality have largely produced estimates for the year 2020,^3–6,24^ leaving patterns of excess mortality during 2021 and 2022 under-studied. The Center for Disease Control and Prevention (CDC) has reported an estimate of approximately 1.1 million excess deaths in the U.S. from March 2020 to February 2022, which is in line with our estimate.^25^

Generating estimates of excess mortality at the county-level has several potential benefits. First, because counties are the administrative unit for death investigation, excess mortality estimates have the potential to help identify counties where Covid-19 death rates differ from excess mortality rates and who might benefit from additional training in cause-of-death certification.^26^ Such estimates may also be valuable for informing local public health workers, community leaders, and residents of the true impact of the pandemic, thus potentially increasing vaccination and uptake of other protective measures.^27^ These estimates may also be used to target federal and state emergency resources, such as funeral assistance support from the Federal Emergency Management Agency (FEMA). Finally, estimating excess mortality at the county-level also enables analyses of social and structural factors affecting mortality associated with the pandemic, including across geographic dimensions like metropolitan status.

One major finding of this study is that the number of excess deaths in the second year of the pandemic was not substantially lower than the first year, which is noteworthy as vaccinations were available for much of 2021 and 2022. Despite the strong efficacy of vaccines, gaps in uptake likely contributed to high excess mortality in 2021 and 2022, which may persist into the future if these vaccination gaps are not closed. This finding may also reflect federal and state governments’ failure to invest in population-based strategies designed to protect the communities at greatest risk for Covid-19 death, such as financial support for family and medical leave, improved ventilation of schools and workplaces, and vaccine delivery programs organized in coordination with community partners.^28^

A second and related major finding of this study is that excess mortality moved substantially from large metro areas in the first year of the pandemic to nonmetro areas during the second year. One factor that likely contributed to this change is vaccination. In urban areas, 75% of people aged 5 years and older were vaccinated as of January 2022 compared to only 59% of people aged 5 years and older in rural areas.^29,30^ This rural-urban difference in vaccination rates has more than doubled since April 2021, suggesting that differences in vaccination rates across metro-nonmetro categories may be playing an increasingly important role in the rural mortality disadvantage observed in the second year of the pandemic. A prior study found that increases in Covid-19 mortality in nonmetro areas during the second year of the pandemic were largely driven by increases in mortality among non-Hispanic white populations.^31^ In line with this, a survey from 2021 found that white, older individuals who identified as Republican were the most vaccine hesitant population in the U.S..^32^ Another factor that may be contributing to high rural excess mortality is insufficient rural health infrastructure related to funding gaps and workforce shortages.^33^ This may have affected access to Covid-19 treatment, including oral antivirals and monoclonal antibody treatments.^34^ Another consideration is the high prevalence of comorbidities among rural residents that likely increased risk for severe Covid-19 outcomes.^35^ Each of these factors may have contributed to the rural mortality disadvantage observed in this study.

The study had several limitations. First, the study relied on publicly available data, which were subject to suppression of death counts fewer than 10 in a given county-month. We addressed this limitation by pooling information across different geographical levels through the use of hierarchical models and by taking advantage of the additional information provided by yearly death counts. However, our estimates remain uncertain in areas with small populations and few deaths. Second, our study examined all-cause mortality and did not explore differences in trends using cause-specific death rates. Assessing geographic and temporal differences in excess death rates by cause-of-death would allow for a deeper understanding of the mechanisms driving trends in excess mortality overall and is an important direction for future work. Third, we were not able to model age-specific excess mortality at the county-month level because of suppression in CDC public data. However, the number of excess deaths across all ages combined is an important metric of the impact of the pandemic in a given area even when its magnitude is partially explained by age distribution. Finally, the primary objective of the present study was to generate descriptive estimates of excess mortality for each county over the course of the pandemic. As such, we did not model the determinants of spatial-temporal variation in excess mortality. An important direction for future research will be to identify the key demographic, social, structural, and policy factors that contributed to differences in county excess mortality across time and space to gain insight into why some counties experienced much worse outcomes than others.

In conclusion, this study provides the first county-level estimates of excess mortality by month in the U.S. during the first two years of the pandemic (March 2020 to February 2022). It reveals that the burden of excess mortality has moved substantially from large metro areas in the first year to nonmetro areas in the second year, especially in the South. Future research should use the estimates generated here to examine the social and structural factors associated with excess mortality throughout the pandemic, counties where Covid-19 death rates differ significantly from excess death rates, and mechanisms contributing to rural health disparities during the pandemic.

## Methods

### Data

Monthly death counts at the county level were extracted from the CDC WONDER online tool. See **Methods Supplement** for further details about data extraction procedures. We extracted all-cause death counts from the Multiple Cause of Death database using the provisional counts for 2021 and 2022 and the final counts for 2015-2020. To convert the number of deaths into rates, we used publicly available yearly county-level population estimates from the Census Bureau (2010-2020^36^ and 2021^37^). To compute monthly rates, we assumed linear growth in population between each two time points. For the August 2021-February 2022 period, for which no population estimates are available, we projected county-level population by applying the county-specific average monthly growth rate for the period January 2018 - July 2021 (the most recent month for which Census Bureau estimates were available).

We harmonized county FIPS codes by reversing FIPS code changes implemented by the Census Bureau (code changes, merging of counties, or separation of counties) until we could ensure that FIPS code represented the same spatial units across all data sources.^38^ This harmonization procedure led to a total of 3,127 units. For exploration of the results of our model, we grouped counties into 4 metropolitan-nonmetropolitan categories (large central metro, large fringe metro, medium or small metro, and nonmetro) based on the 2013 NCHS Rural-Urban Classification Scheme for Counties.^39^ For simplicity of comparison, in some analyses, we reduced the 4 metropolitan-nonmetropolitan categories into 2 or 3 groups. We also grouped counties into 4 Census Bureau Regions (Northeast, Midwest, South, and West) and 9 Census Bureau Divisions (New England, Middle Atlantic, East North Central, West North Central, South Atlantic, East South Central, West South Central, Mountain, and Pacific). Finally, in some analyses, we stratified the Census Regions and Divisions by the metropolitan-nonmetropolitan categories, leading to more granular geographic units. See **Methods Supplement** for further details about the geographic classifications used in this study.

Within the two-year period, we identified four temporal peaks representing periods where Covid-19 mortality in the U.S. was heightened. We identified peaks as periods where excess death rates rose steeply and then steeply declined. Four peaks were identified: the initial wave (March to August 2020), the winter wave (October 2020 to February 2021), the delta wave (August to October 2021) and the omicron wave (November 2021 to February 2022).

### Statistical Methods

To predict the monthly county-level number of deaths, we fit a Bayesian hierarchical model starting from the framework described in a prior paper^22^ and adapting it to our specific application. Let *y*_*ts*_ be the number of deaths in spatial unit *s* at time *t*. Let *P*_*ts*_ be the population of spatial unit *s* at time *t*. We assume a Poisson distribution for the number of monthly deaths *y*_*ts*_ and model the risk *r*_*ts*_ of dying using the following specification:

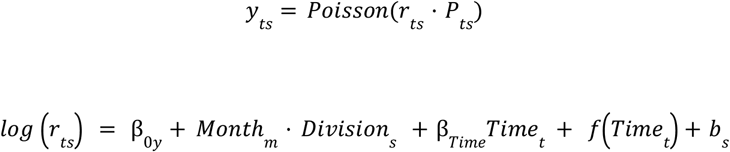

where β_0*y*_ is the year specific intercept given by β_0*y*_ = β_0_ + ε_*y*_, with β_0_ being the global intercept and 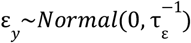 an unstructured random effect representing the deviation of each year from the global intercept. The parameter τ_ε_ indicates the precision of ε_*t*_. We include fixed effects for each month and Census division to capture seasonal effects. The linear predictor also includes both a linear effect (captured by β_*Time*_) and non-linear effect *f*(⋅) of time (in months) since the start of the period (*t* = 1, 2, … with time 1 corresponding to January 2015).

For the non-linear effect, we assume the following first-order autoregressive process (AR1) model:

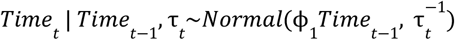

We model county-level intercepts using the modified Besag, York and Mollie spatial model proposed by Riebler et al. (BYM2 model).^40^ This model is the sum of a spatially unstructured random effect,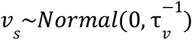 and spatially structured effects μ _*s*_. *b*_*s*_ is defined as:

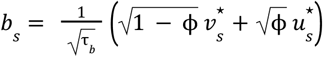

where 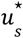 and 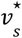 are standardized versions of μ_*s*_ and υ_*s*_ to have variance equal to 1. The term 0 ≤ ϕ ≤ 1 is a mixing parameter which measures the proportion of the marginal variance explained by the spatially structured effect.

We specify minimally informative prior distributions for the fixed effects β_0_, the month-division specific intercepts *Month*_*m*_ *Di*υ*ision m*_*s*_ = 1, 2, …, 12, the linear time effect β_*Time*_, and the ϕ_1_ parameter for the AR1 process. For the hyperparameters of the BYM2 model, ϕ and τ_*b*_, we adopt priors that tend to regularize inference while not providing too strong information, the so-called penalized complexity (PC) priors introduced by Simpson et al.^41^ In particular, for the standard deviation 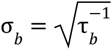 I select a prior so that *Pr* (σ_*b*_ > 1) = 0. 01, implying that it is unlikely to have a spatial relative risk higher than *exp*(2) based solely on spatial or temporal variation. For ϕ we set pr *Pr* (ϕ < 0. 5) = 0. 5 reflecting our lack of knowledge about which spatial component, the unstructured or structured, should dominate the spatial term *b*_*s*_. Finally, we also adopt PC priors for all the remaining standard deviations of random effects 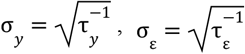, and 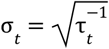 such that for each hyperparameter *Pr* (σ > 1) = 0. 01.

We fit the models using the Integrated Nested Laplace Approximation (INLA) method, through the R-INLA software package.^42^ We train the model on the years 2015–2019. We experimented with a longer training window (2010-2019) but found no meaningful improvements in performance with respect to our final choice.

### Suppressed Observations

Death counts less than 10 were censored in the public data used for analysis. Between 2015-2019, 1,312 distinct counties had at least one month of censored data, totaling 42,734 county-months. To address suppression of death counts below 10, we estimated censored death counts with a set of state-year specific censored Poisson models using monthly dummies to capture seasonality, and imputed the suppressed observations with the estimated counts. We exploited lower levels of censoring in year-level data to further adjust the total of imputed deaths by year and state to sum to the difference between the total of uncensored month-level deaths aggregated to the year level and the uncensored year total (obtained from a year-level data extract).

### Model Validation

We performed a cross-validation procedure to evaluate the out-of-sample validity of the predictions generated by our methods. Using data for the years 2015–2019, we fit the proposed model multiple times on data from a random sample of states. After training the model on 2015 data and predicting 2016-2019 death counts, we then trained the model on 2015-2016 data and predicted 2017-2019 death counts, and so on. We assessed the agreement between the predicted and observed deaths in the year(s) excluded from the training data and average over the cross-validation results using the following metrics: a) the correlation between predicted and observed deaths and b) 90% coverage, defined as the probability that the observed deaths lie within the 90% credible interval estimated from the model. Results from this cross-validation procedure, stratified by metro category and Census region, are presented in **Supplementary Table 1**. All strata achieved correlation > .89 and 90% coverage > .90, with the vast majority > .95 and > .93, respectively. Sample output for the largest counties in each Census Division and Metro Category are provided in **Supplementary Table 2**.

This study used de-identified publicly available data and was exempted from review by the Boston University Medical Center Institutional Review Board. Programming code was developed using R, version 4.1.0 (R Project for Statistical Computing) and Python, version 3.7.13 (Python Software Foundation).

## Data Availability

Data used in the study are publicly available from the US Centers for Disease Control and Prevention and US Census Bureau. Additional details about the data and programming code for replication can be accessed at the linked GitHub repository: https://github.com/The-Uncounted-Lab/county_month_excess_mortality_2020-2022

https://mu0brt-zhenwei-zhou.shinyapps.io/county_ex_app/

## Acknowledgements

The authors would like to thank Robert Anderson and Farida Ahmad (National Center for Health Statistics) for providing assistance with the provisional mortality files and Marta Blangiardo (Department of Epidemiology and Biostatistics, School of Public Health, Imperial College London) for input on the statistical modeling. Additionally, the authors would like to thank Ahyoung Cho, Steele Myrick, Mikas Hansen, Sylvia Lutze, and Elif Coskun for technical and administrative support.

## Supplementary Online Content

## Supplementary Methods

**Supplementary Multimedia**. RShiny App. Interactive Map of Monthly Excess Mortality Estimates for 3,127 Counties. https://mu0brt-zhenwei-zhou.shinyapps.io/county_ex_app/

## Methods Supplement

### Data Extraction from CDC WONDER

The CDC WONDER online database query system found at https://wonder.cdc.gov/ was used to extract all mortality data used in this project. To obtain death counts for all-causes mortality, we used the Multiple Cause of Death (Final) database from 1999-2020. We obtained two main sets of extracts, one for data at the county-month level and one for data at the county-year level (to investigate suppression). In order to minimize suppression, additional extracts were obtained at the wave level and the pandemic year level (pooling different months) for all figures and tables using these longer time periods. For all cause mortality extracts, the data request was submitted for the time period of interest using the request form with the following settings:

- Tab 1. Organize table layout: Group results by County and by: Month for monthly data, Year for yearly data, and no additional group for wave and pandemic year data
- Tab 4. Select time period of death: specific period
- Tab 6. Select underlying cause of death: *All* (All Causes of Death)
- Tab 8. Other options: checking Export Results. The request generates a text file.

To extract data for the time periods of March 2020 to February 2022, we used the Multiple Cause of Death (Provisional) database from 2018 – Last Month database. The data requests were submitted for each time period of interest using the request form with the following settings:

- Tab 1. Organize table layout: Group results by County and by: Month for monthly data, Year for yearly data, and no additional group for wave and pandemic year data
- Tab 4. Select time period of death: March 2020 to February 2022
- Tab 6. Select underlying cause of death: *All* (All Causes of Death)
- Tab 8. Other options: checking Export Results. The request generates a text file.

### Geographic Classifications

USDA/ERS/NCHS Metropolitan-Nonmetropolitan Categories:

- **Large central metros**: counties in metropolitan statistical areas with a population of more than 1 million.
- **Large fringe metros**: counties that surrounded the large central metros
- **Small or medium metros**: counties in metropolitan statistical areas with a population between 50,000 and 999,999.
- **Nonmetropolitan areas**: all other counties.

US Census Divisions:

- **New England**: Connecticut, Maine, Massachusetts, New Hampshire, Rhode Island, Vermont
- **Middle Atlantic**: New Jersey, New York, Pennsylvania
- **East North Central**: Indiana, Illinois, Michigan, Ohio, Wisconsin
- **West North Central**: Iowa, Kansas, Minnesota, Missouri, Nebraska, North Dakota, South Dakota
- **South Atlantic**: Delaware, District of Columbia, Florida, Georgia, Maryland, North Carolina, South Carolina, Virginia, West Virginia
- **East South Central**: Alabama, Kentucky, Mississippi, Tennessee
- **West South Central**: Arkansas, Louisiana, Oklahoma, Texas
- **Mountain**: Arizona, Colorado, Idaho, New Mexico, Montana, Utah, Nevada, Wyoming
- **Pacific**: Alaska, California, Hawaii, Oregon, and Washington US Census Regions:
- **Midwest:** Indiana, Illinois, Michigan, Ohio, Wisconsin, Iowa, Kansas, Minnesota, Missouri, Nebraska, North Dakota, South Dakota
- **Northeast:** Connecticut, Maine, Massachusetts, New Hampshire, Rhode Island, Vermont, New Jersey, New York, Pennsylvania
- **South:** Delaware, District of Columbia, Florida, Georgia, Maryland, North Carolina, South Carolina, Virginia, West Virginia, Alabama, Kentucky, Mississippi, Tennessee, Arkansas, Louisiana, Oklahoma, Texas
- **West:** Arizona, Colorado, Idaho, New Mexico, Montana, Utah, Nevada, Wyoming, Alaska, California, Hawaii, Oregon, and Washington

### Model Definition in INLA

We report here the model definition in INLA. This should help the reader understand the specifics of the model and interpret Supplementary Figures 2, 3, and 4, and Supplementary Table 2, reporting (graphically or in table format) the posterior distribution of the model’s parameters and hyperparameters with the exception of the random effects for the counties.

~~~
*# priors
hyper.bym <- list(theta1 = list(‘PCprior’, param=c(1, 0.01)),
theta2 = list(‘PCprior’, param=c(0.5, 0.5)))
hyper.iid <- list(theta = list(prior=“pc.prec”, param=c(1, 0.01)))
hyper.ar1 <- list(theta1 = list(prior=“pc.prec”, param=c(1, 0.01)))
# model formula
formula <- deaths ∼ 1 + offset(log(pop)) + timeID +
as.factor(month)*as.factor(divisionID) +
f(FIPSID, model=‘bym2’, hyper=hyper.bym,
 graph=inla.graph, scale.model=T) +
f(yearID, model=‘iid’,hyper=hyper.iid) +
f(timeID2, model=‘ar’,order=p,hyper=hyper.ar1)*
~~~

**Supplementary Table 1.**
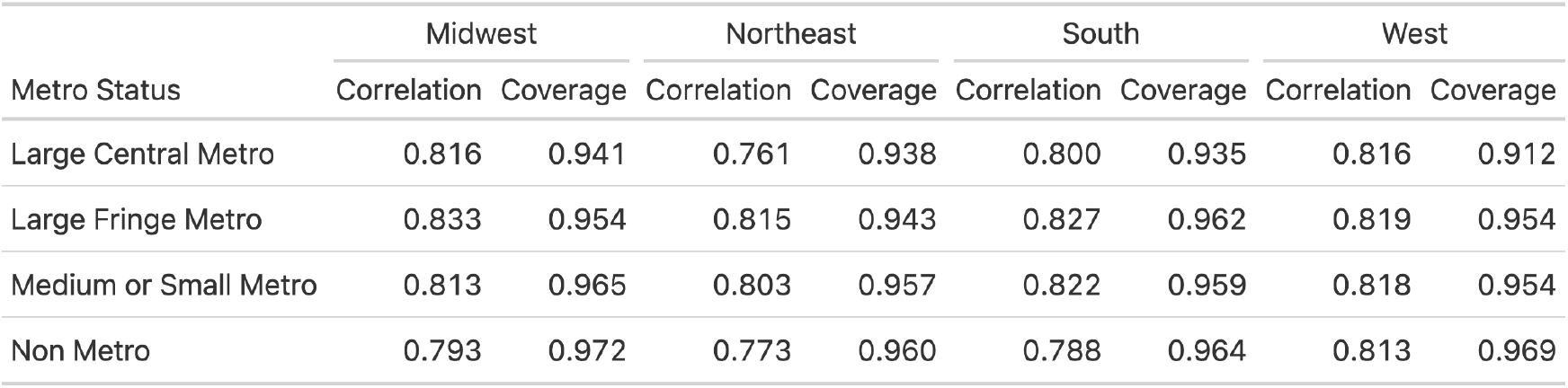
Cross-validation results for the model for monthly death counts: correlation between predicted and observed number of deaths, and 90% coverage probability across the different metro/non-metro categories and Census Regions.

| Metro Status | Midwest |  | Northeast |  | South |  | West |  |
| --- | --- | --- | --- | --- | --- | --- | --- | --- |
|  | Correlation | Coverage | Correlation | Coverage | Correlation | Coverage | Correlation | Coverage |
| Large Central Metro | 0.816 | 0.941 | 0.761 | 0.938 | 0.800 | 0.935 | 0.816 | 0.912 |
| Large Fringe Metro | 0.833 | 0.954 | 0.815 | 0.943 | 0.827 | 0.962 | 0.819 | 0.954 |
| Medium or Small Metro | 0.813 | 0.965 | 0.803 | 0.957 | 0.822 | 0.959 | 0.818 | 0.954 |
| Non Metro | 0.793 | 0.972 | 0.773 | 0.960 | 0.788 | 0.964 | 0.813 | 0.969 |

**Supplementary Table 2.** Summary of the Posterior Distribution for the Model’s Hyperparameters and for the Linear Time Trend

|  | Mean | S.D. | Percentile |  |  |
| --- | --- | --- | --- | --- | --- |
|  |  |  | 2.5th | 50th | 97.5th |
| Precision for FIPSID | 11.1780 | 0.4381 | 10.3794 | 11.1546 | 12.1027 |
| Phi for FIPSID | 0.8749 | 0.0151 | 0.8429 | 0.8757 | 0.9023 |
| Precision for yearID | 1,939.6508 | 1,153.7022 | 681.5905 | 1,626.5955 | 4,993.6974 |
| Precision for timeID2 | 500.4373 | 178.4131 | 272.2303 | 459.8157 | 953.2002 |
| Rho for timeID2 | 0.9320 | 0.0132 | 0.9043 | 0.9326 | 0.9557 |
| timeID | 0.0004 | 0.0013 | -0.0021 | 0.0004 | 0.0029 |

**Supplementary Figure 1.**
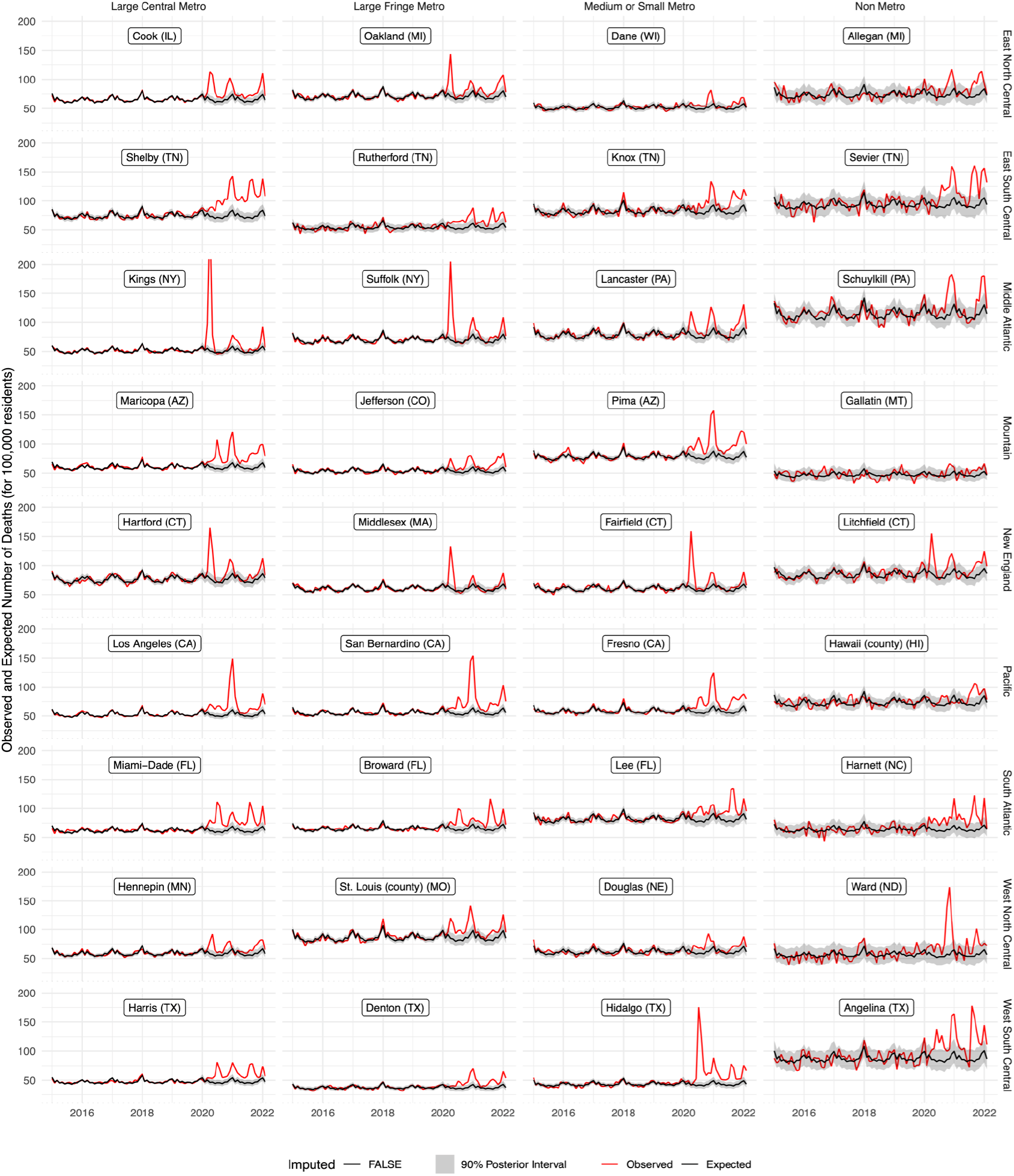
Sample Model Output for the Largest Counties in each Census Division and Metro Category Notes: Each facet represents observed (red) versus expected (black) deaths for 100,000 residents. The gray area around the line for expected deaths represents 90% the posterior interval. Counties are aligned on a grid where rows represent Census Divisions and columns metro categories. Counties were selected to be the largest (by population) in each cell.

**Supplementary Figure 2.**
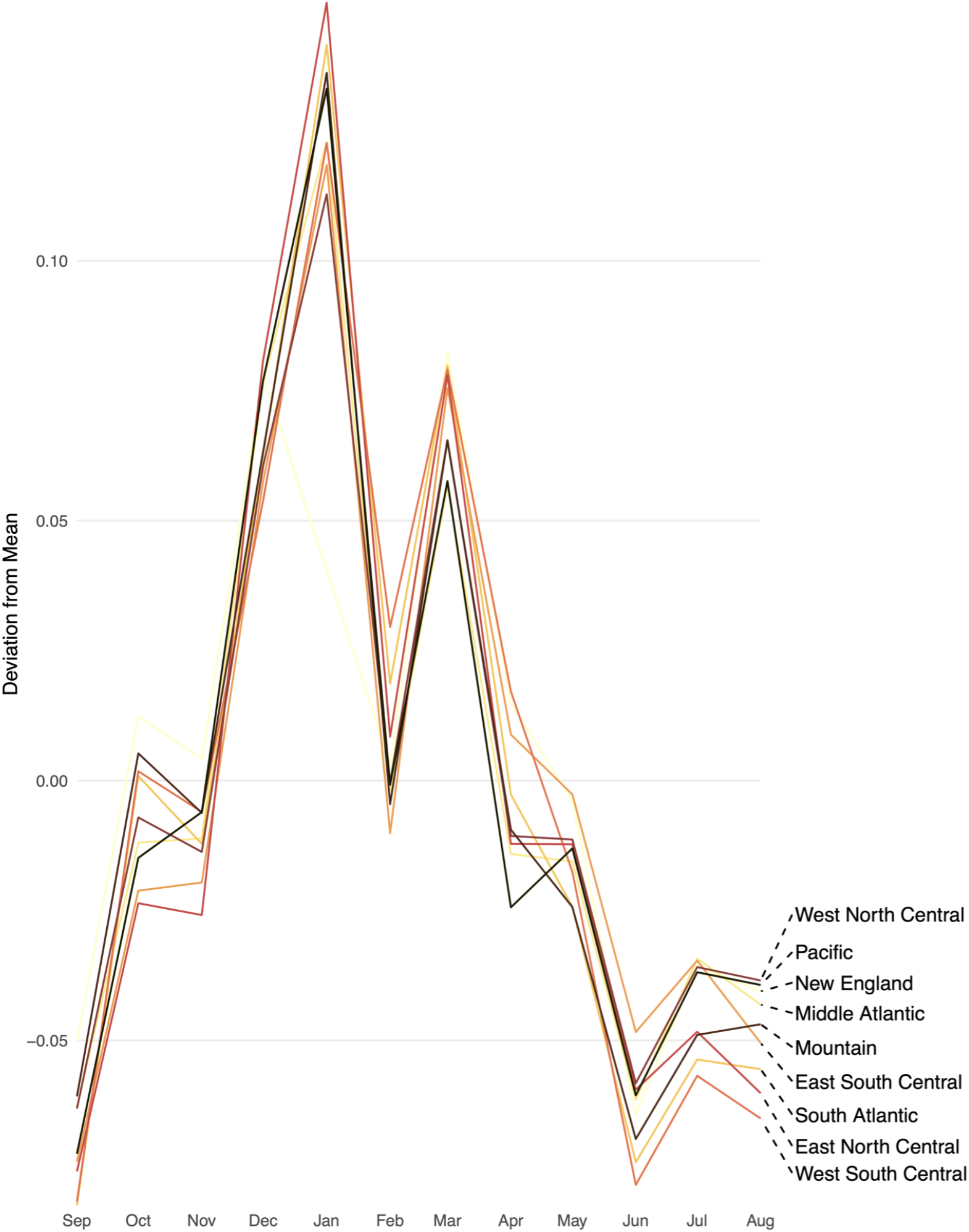
Month seasonal effects by Census Division

**Supplementary Figure 3.**
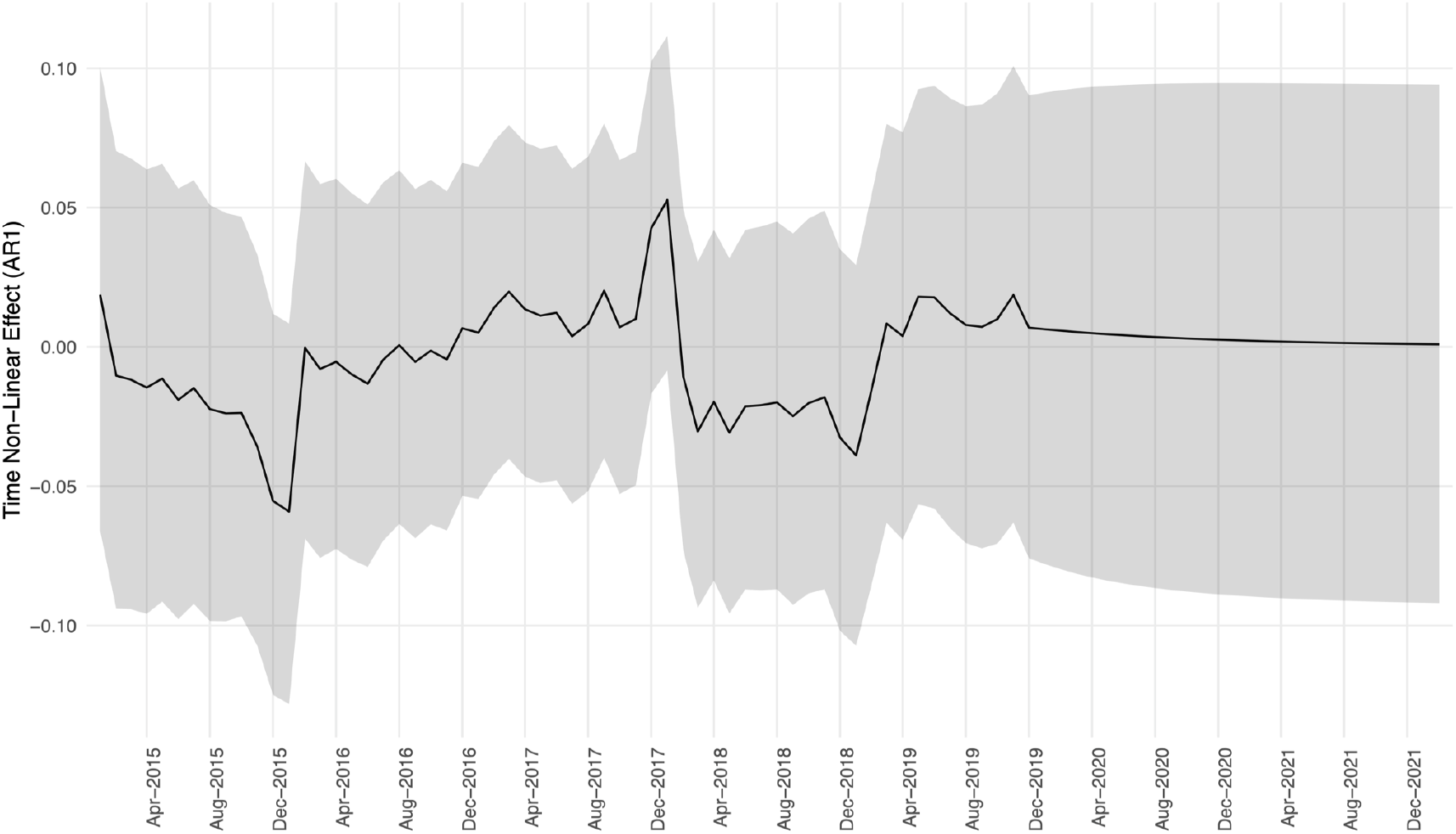
Non-Linear Effect of Time (AR1 Component) with 95% Posterior Interval

**Supplementary Figure 4.**
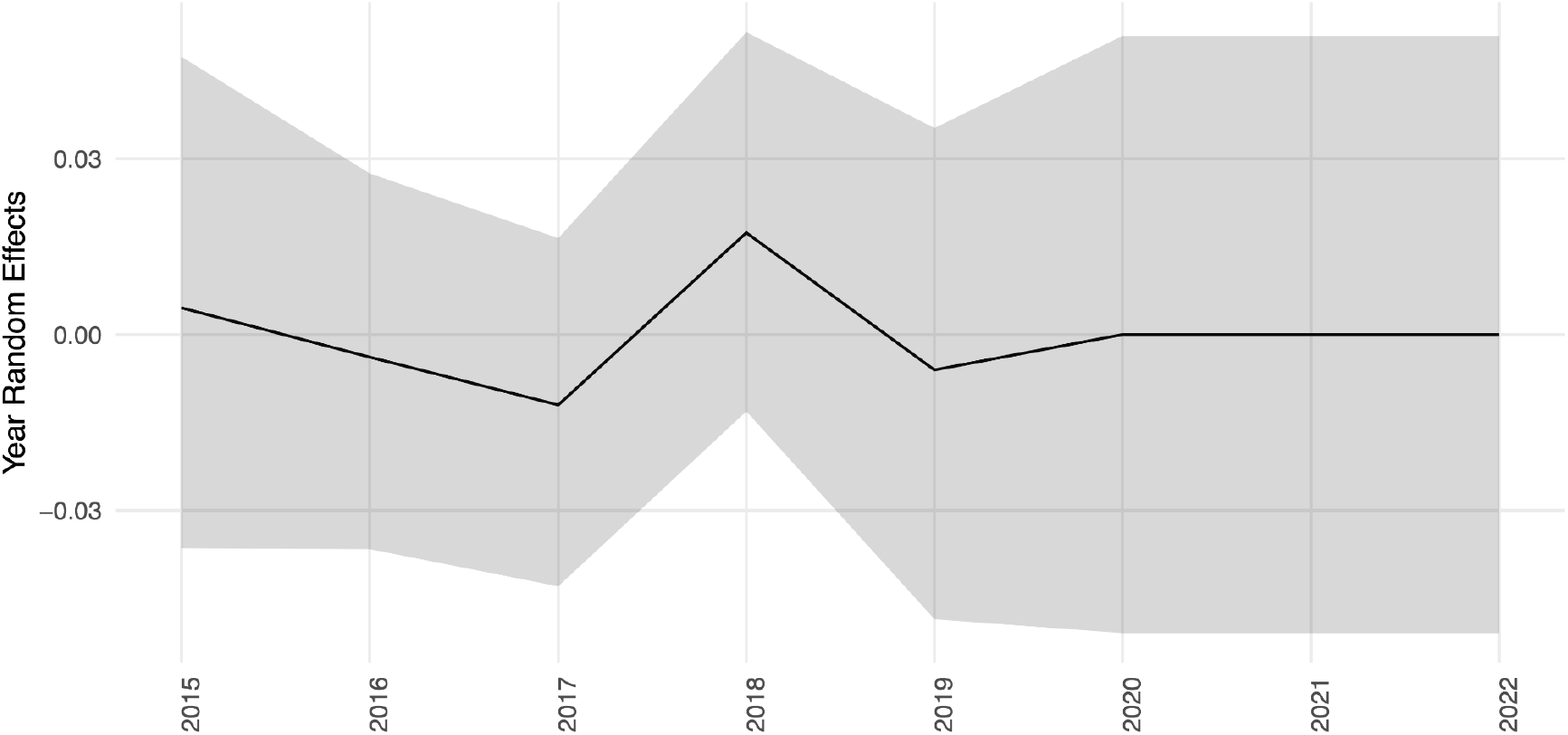
Year Random Effects with 95% Posterior Interval

